# OUTCOME OF COVID-19 PATIENTS ON STEROID THERAPY: A LONGITUDINAL STUDY

**DOI:** 10.1101/2023.08.03.23293425

**Authors:** Anant Kataria, Prasan Panda, Yogesh Bahurupi, Gaurav Chikara

**Affiliations:** Department of Internal medicine, All India Institute of medical science, Rishikesh; Department of family and community medicine, All India of medical science, Rishikesh; Department of Pharmacology, All India Institute of medical science Rishikesh

## Abstract

**Introduction:** SARS-CoV-2 is responsible for global pandemic that originates from Wuhan, China (1). Patients’ presentation van be varied from asymptomatic to severe ARDS and multiorgan dysfunction likely due the dysregulated systemic inflammation (2). Glucocorticoids inhibits the inflammation by down streaming of cytokine receptor and promote resolution (3). The role of corticosteroid in COVID-19 still remains controversial. Corticosteroids associated with many long terms side effects. Previous MARS outbreak had experienced avascular necrosis with corticosteroid use (4).

**Objectives:** The aim of the study was to evaluate the outcome of covid-19 patients on the corticosteroid therapy and estimate mortality rate with corticosteroid therapy and investigate potential long-term adverse events associated with its use.

**Methods:** We did a longitudinal follow up study at the AIIMS Rishikesh to assess the side effects of corticosteroids in COVID-19 patients. Patients with moderate to severe COVID-19 pneumonia requiring the oxygen support were included in the study. According to the institutional protocol patients received conventional dose steroids versus pulse dose steroids. (Based on CT/ X-ray findings). Patients were followed up in the hospital till discharge/death for assessment of adverse events due to corticosteroids and all other biochemical parameters (Inflammatory markers) and SOFA score were obtained during hospitalisation till discharge. And at the 6 month follow up patient was assessed for infection and avascular necrosis of the femur.

**Results:** A total of 600 patients were screened out of which 541 patients who received corticosteroids were included in this study. 71.3% were male and 26.6 % were females. Most prevalent comorbidity was systemic hypertension (38.8%) followed by diabetes mellitus (38%). Most common presenting symptoms was dyspnoea followed by fever and cough. Majority patients received dexamethasone (95%). 65.8 % patients received conventional dose while 34.2% of patients received pulse dose. Mortality was more associated with pulse dose (44%) then a conventional dose (30%) (p-value 0.0015). the median duration of the corticosteroids was 10 days with an IQR of 7-13 days. During the hospitalisation 142 patients (26.2%) develops hyperglycaemia. Hyperglycaemia was more prevalent in the pulse dose steroid group (16.8% versus 9.4%). One patient develops pancreatitis. There was a significant reduction in the levels of inflammatory markers (p<0.005) after steroid initiation. At the 6th month of follow patients were assessed for AVN and suspected infection. 25 patients (8.25%) had infection out of which 19 received pulse dose. Out of 25 patients’ cultures was available for 7 patients and 2 patients grows pathogenic organism in the urine (pseudomonas and E. coli). 02 patients develop non-specific joint pain at 6 months. No patient had AVN during the follow up.

**Conclusion:** Corticosteroid therapy in the COVID-19 is associated with various adverse event, commonly hyperglycaemia and the risk of the same increased with the high dose corticosteroids. corticosteroids appear to be a double-edged sword in combat against COVID-19 and need to be used aptly considering the risk-benefit ratio. The outcome of COVID-19 patients on corticosteroid therapy varies due to the use of different doses of corticosteroids. Routine follow-up of the recovered patients is needed to detect early unwanted

## INTRODUCTION

Coronavirus disease 2019 (COVID-19) caused by severe acute respiratory syndrome coronavirus 2 (SARS-CoV-2) is responsible for the global pandemic that originates in Wuhan, China in December 2019. Acute respiratory distress syndrome (ARDS) usually develops from the second week onwards (1). Approximately 10% require hospital admission because of Covid-19 pneumonia, approximately 10% of which require Intensive Care Unit (ICU) care, including some who need invasive ventilation due to acute respiratory distress syndrome (ARDS) (2). There is no specific anti-viral drug regime used to treat critically ill patients. The management of patients mainly focuses on the provision of supportive care with oxygenation, ventilation, and fluid management. Combination treatment of low-dose systematic corticosteroids and anti-viral has been encouraged as part of critical COVID-19 management (3). Most of the earlier studies done on severe acute respiratory syndrome 1 (SARS-CoV-1) and Middle Eastern Respiratory Syndrome (MERS-CoV) showed adverse outcomes with corticosteroid treatment (4). In many guidelines for the treatment of COVID-19, patients have stated that glucocorticoids were either contraindicated or not recommended (5). The practice has varied across the world, as many as 50 % of patients have been treated with corticosteroids (6) (7). There are few studies on the dose of corticosteroids in COVID-19. The use of pulse doses of steroids has been studied during SARS and MERS pandemics with the results being controversial (8). Upcoming studies showed that pulse steroid therapy reduces the progression of COVID-19(9) (10). Corticosteroids are associated with many short-term side effects like hyperglycaemia, cutaneous manifestations, electrolyte abnormalities, hypertension, pancreatitis, neuropsychological effects, and hematologic and immunologic effects. Long-term effects include aseptic joint necrosis, osteoporosis, adrenal insufficiency, and gastrointestinal, ophthalmic, and hepatic effects (11)

This study explores the outcome of COVID-19 patients on corticosteroid therapy and the long-term adverse effect associated with use of it. We have analysed the data from the patients admitted in the tertiary care COVID-19 hospital from June 2020 to May 2021.

## METHADOLOGY

The study was carried out at All India Institute of Medical Sciences (AIIMS), Rishikesh, a tertiary care centre of North India’s population. The data was obtained from the hospital database system and analysed the records of lab confirmed COVID-19 patients of at least 18 years of the age. The biochemical investigations were performed according to the institutional protocol. Complications during the hospitalization-shock, cardiac arrhythmias, cardiac arrest, pulmonary embolism/pneumomediastinum, myocarditis, pancreatitis, acute kidney injury, and hyperglycaemia were obtained. Discharged patients were followed up at 06 months. Rehospitalisation and mortality were obtained telephonically or in the OPD. For assessment of long-term side effects of corticosteroids (a secondary bacterial infection and avascular necrosis of the head of the femur), if the suspected infection-total leucocyte counts, blood, and urine culture were obtained, and for avascular necrosis of the head of the femur hip, X-ray was obtained. Patients were started on the corticosteroids therapy according to the institutional protocol.

## STUDY FLOWCHART (figure 1)

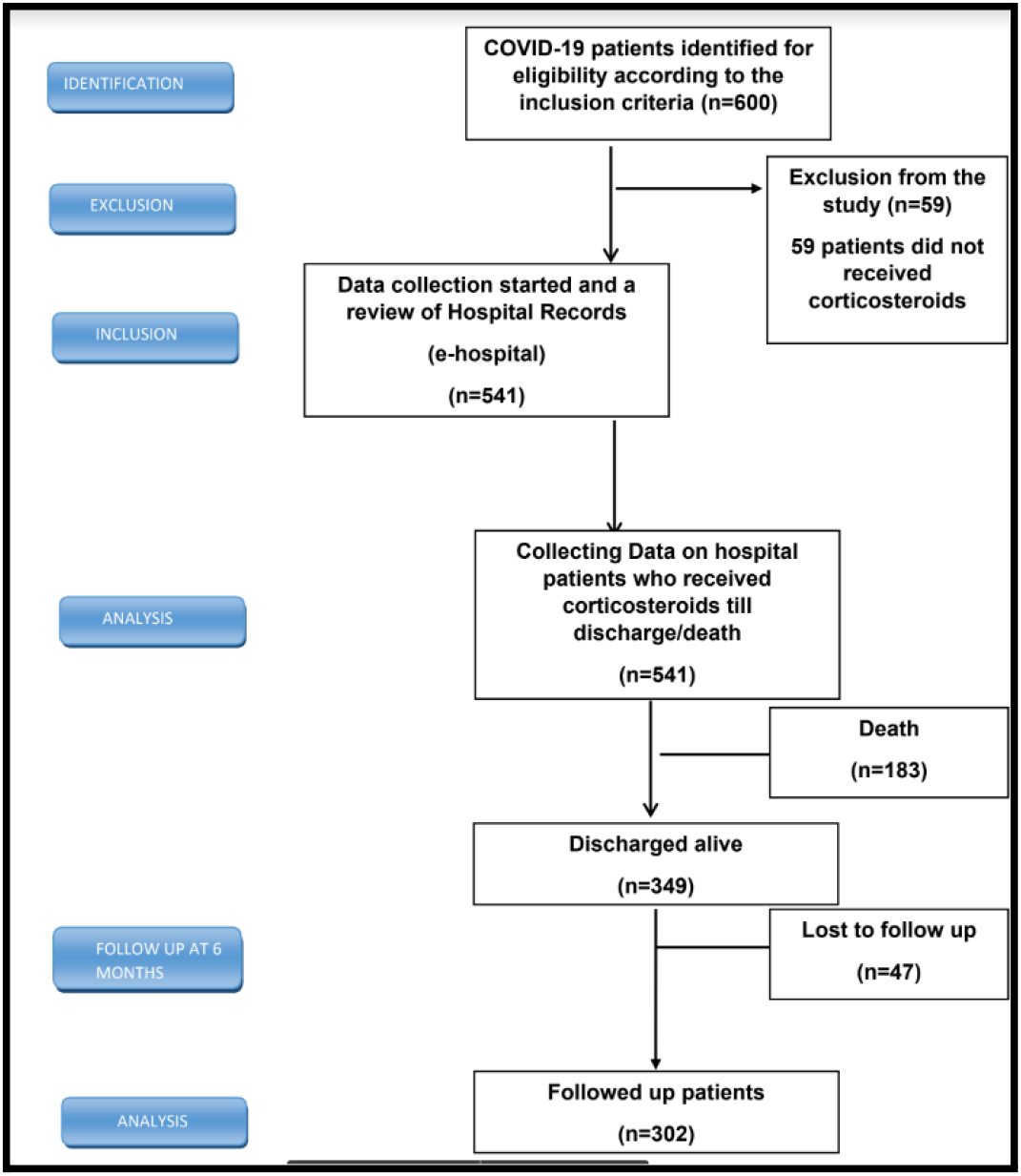

## STATISTICAL ANALYSIS

Categorical variables were presented in number and percentage (%), and continuous variables were presented as mean ± SD and median, median (IQR). The normality of the data was tested by the Kolmogorov-Smirnov test. If the normality was rejected, then a non-parametric test was used.

1. Qualitative variables were correlated using the Chi-square test.
2. Qualitative variables were compared using the independent t-test between the two groups for >2 groups, analysis of variance (ANOVA) was used.
3. Wilcoxon-Mann-Whitney test was used to compare the two arms (non-parametric i.e., comparison of ICU stay, oxygen requirement.
4. Friedman test was used for the detection of changes in the SOFA score and inflammatory markers.

A p-value of <0.05 was considered statistically significant.

The data was entered in the Microsoft EXCEL spreadsheet, and analysis was done using IBM Statistical Package for Social Sciences (SPSS) version 26.0 (Chicago, US).

## RESULTS

Between June 2020 to May 2021, we analysed records of 541 patients (figure 1). 68.7% patients were between 18 to 60 years of age group while 31.2 % patients were more than 60 years of age (table 1). 71.3 % were males and 26 % patients were females (table 2, figure 2). Out of 541 patients, 183 patients died (mortality rate 33.64%) and nine patients (1.66%) were transferred to another hospital / leave against medical advice.

**Table1:**
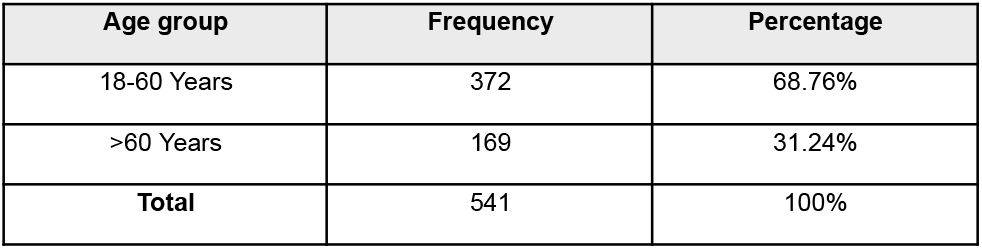
Distribution of participants in terms of Age (N = 541)

**Table 2:** Distribution of participants in terms of Gender (N = 541)

| Gender | Frequency | Percentage |
| --- | --- | --- |
| Male | 386 | 71.34% |
| Female | 155 | 28.66% |
| **Total** | 541 | 100 % |

**Figure 2:**
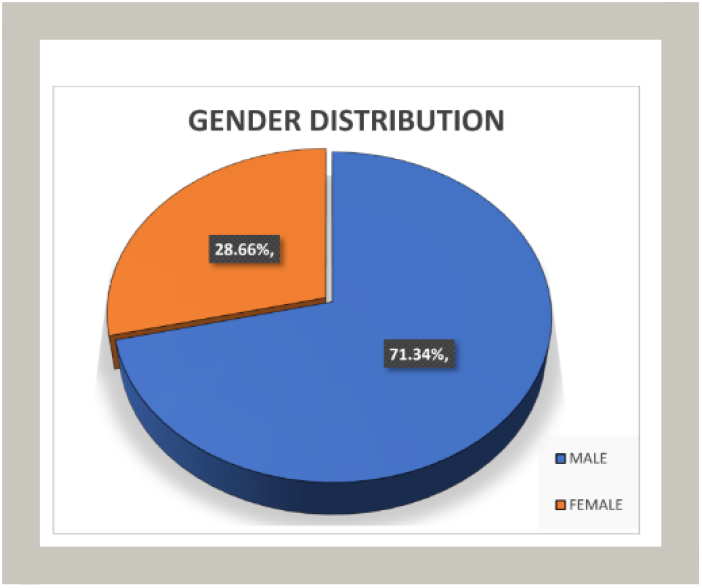
Distribution of participants in terms of Gender (N = 541)

Majority patients received dexamethasone (95%). 65.8 % patients received conventional dose while 34.2% of patients received pulse dose (Table 5). Mortality was more associated with pulse dose (44%) then a conventional dose (30%) (P-value 0.0015). The median duration of the corticosteroids was 10 days with an IQR of 7-13 days. During the hospitalisation 142 patients (26.2%) develops hyperglycaemia. Hyperglycaemia was more prevalent in the pulse dose steroid group (16.8% versus 9.4%). There was a significant reduction in the levels of inflammatory markers (p<0.005) after steroid initiation.

**Table 3:** Co-morbidities existing at admission (N = 541)

| Pre-existing co-morbidity | Frequency | Percentage |
| --- | --- | --- |
| Systemic hypertension | 210 | 38.8% |
| Diabetes mellitus | 206 | 38.07% |
| Chronic obstructive pulmonary disease | 38 | 7.02% |

**Table 4:** symptoms at admission (N = 541)

| Symptom | Frequency | Percentage |
| --- | --- | --- |
| Fever | 437 | 80.77% |
| Cough | 412 | 76.15% |
| Dyspnea | 440 | 81.33% |
| Chest pain | 28 | 5.10% |
| Sore throat | 40 | 7.39% |
| Conjunctivitis | 5 | 0.9% |
| Myalgia | 41 | 7.57% |
| Loss of taste | 7 | 1.30% |
| Loss of smell | 5 | 0.92% |
| Diarrhea | 19 | 3.5 % |

**Table 5:** Descriptive analysis of Corticosteroid therapy (n=541)

| Medication variable | Result |
| --- | --- |
| Dexamethasone, n (%) | 518 (95.74%) |
| Methylprednisolone, n (%) | 22 (4.06%) |
| Prednisolone, n (%) | 01 (0.18 %) |
| Median Duration of Corticosteroids, Days (IQR) |  |
| All patients | 10 (7 to 13) |
| Survivors | 10 (7 to 12.5) |
| Non-survivors | 11 (8 to 13) |
| Dose of Corticosteroids |  |
| Conventional Steroid dose, n (%) | 356 (65.80%) |
| Pulse Steroid dose, n (%) | 185 (34.20%) |
| Median Duration Between the onset of illness and corticosteroid initiation (IQR) | 7 (05 to 09) |
| Median Duration of Clinical recovery after Steroid initiation (IQR) | 10 (08 TO 17.5) |

Time to clinical recovery after steroid initiation was considered with a resolution of signs and symptoms e.g., fever, dyspnoea, and the decrement in the oxygen requirement. The mean time to clinical recovery in the conventional dose corticosteroids was less compared to the pulse corticosteroid group (12.35 days vs 15.46 days, p-value 0.002). There was a statistically significant association in the changes in SOFA score after the of Corticosteroid therapy (p-value 0.026). There was a statistically significant association in the changes in the level of serum procalcitonin (p-value 0.012), CRP (p-value 0.012), serum ferritin (p-value 0.001), d-dimer (p-value <0.0001), serum LDH (p-value <0.0001) after the initiation of the corticosteroid therapy.

The mean duration of oxygen therapy in the conventional dose corticosteroids is less compared to the pulse dose corticosteroids. (12.27 days vs 13.48 days, p-value 0.037). The mean duration of ICU stay in the conventional dose corticosteroids is less compared to the pulse dose corticosteroids. (11.5 days vs 12.63 days, p-value 0.058).At 6 month of follow up 10.26 % patients were found to be dead. Twenty-five patients (8.25%) had suspected infection and 18.8 % patients were rehospitalized. Out of 25 patients with suspected infection, Blood culture and urine culture were obtained from seven patients. Out of seven available cultures, two patients grow pathogenic organisms in the urine (pseudomonas and E. coli), and cultures for five patients were sterile. On telephonic follow-up at 6 months, only two patients complained about hip joint pain. We could not able to obtain their hip joint x-rays.

**Table 6:**
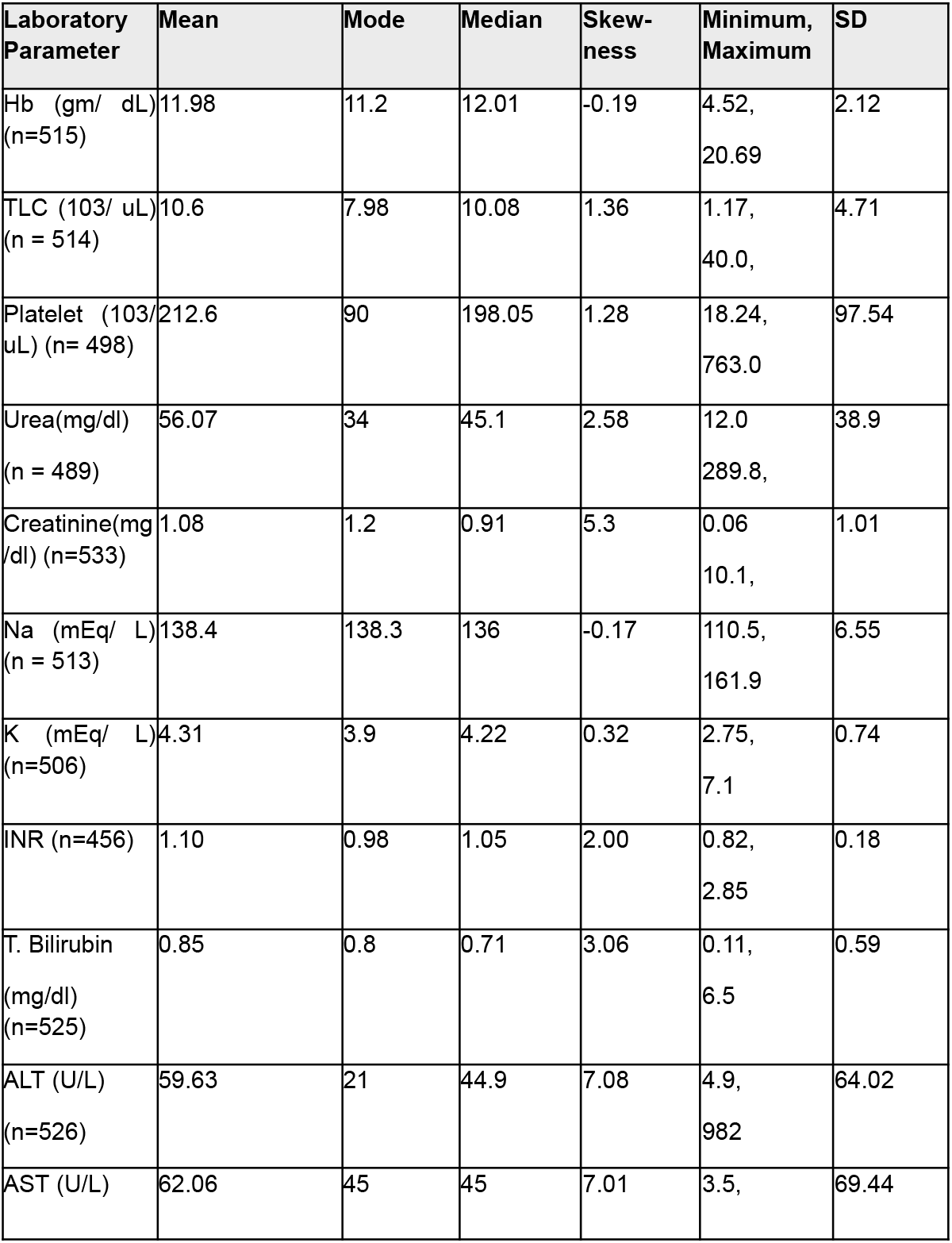
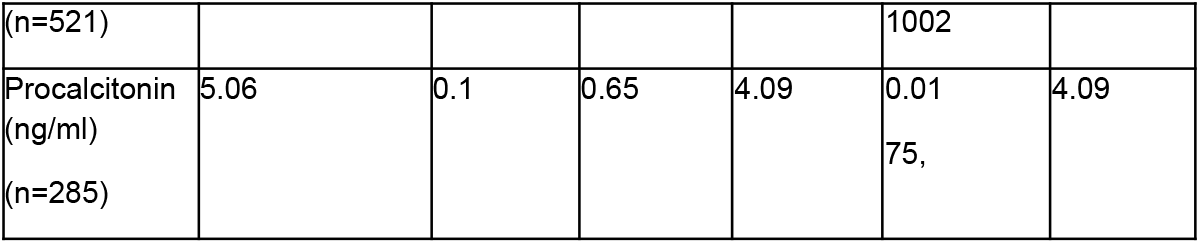
Descriptive statistics of baseline laboratory parameters of study participants.

**Table 7:** Comparison of all-cause mortality between those who received conventional versus pulse doses of corticosteroids.

| Corticosteroids | Final Outcome |  | Total | p-value |
| --- | --- | --- | --- | --- |
|  | Alive | Dead |  |  |
| Conventional Dose | 248(70%) | 105(30%) | 353 | *0.0015 |
| Pulse dose | 101(56%) | 78(44%) | 179 |  |
| **TOTAL** | 349(65%) | 183(35%) | 532 |  |

\*Chi-square test used

**Table 8:** Change in the SOFA score after initiation of corticosteroid therapy:

|  | Mean | SD | p-value |
| --- | --- | --- | --- |
| Baseline SOFA | 3.42 | 1.97 | *0.026 |
| Day 7 SOFA | 3.28 | 3.71 |  |
| Day 14 SOFA | 4.75 | 3.91 |  |
| Day 21 SOFA | 3.06 | 2.48 |  |

\*Friedman test

## DISCUSSION

In a study conducted at the All-India Institute of Medical Sciences Rishikesh, 600 patients with confirmed COVID-19 were admitted between June 2020 and May 2021. Out of these, 541 patients were eligible for the study. The mortality rate among these patients was found to be 33.64%, with a median hospital stay duration of 12 days after starting corticosteroid therapy. The study compared the effects of conventional dose corticosteroids (dexamethasone 6 mg OD or equivalent) and pulse dose corticosteroids (40 mg dexamethasone followed by tapering dose or equivalent). It was observed that the conventional dose was associated with lower mortality (30% vs. 44% for pulse dose). The median duration of corticosteroid therapy was 10 days for both survivors and non-survivors. During hospitalization, 16.8% of patients in the pulse dose group and 9.4% in the conventional dose group developed hyperglycaemia. One patient also developed pancreatitis. At a 6-month follow-up, 8.25% of patients had suspected infection, 18.8% required rehospitalisation, and 10.26% died within 6 months of discharge.

The study also analysed the impact of corticosteroid therapy on various markers and parameters. It was found that there was a statistically significant change in the SOFA score (a measure of organ dysfunction), as well as a significant reduction in inflammatory markers such as d-dimer, ferritin, LDH, procalcitonin, and CRP.Among survivors, the mean duration of oxygen therapy was 11.8 days; with a slightly shorter duration in the conventional dose group compared to the pulse dose group (12.2 days vs. 13.4 days). The mean duration of ICU stay was also shorter in the conventional dose group (11.5 days vs. 12.63 days), although the difference was not statistically significant. Invasive ventilation and the need for inotropic support were associated with higher mortality.

The study referenced other research on corticosteroid therapy in severe COVID-19, which also showed a reduction in mortality. However, it should be noted that the present study did not have a comparison group receiving usual care or placebo. Another study from China found a longer median hospital stay and virus clearance time compared to the present study, which may be attributed to differences in sample size and criteria for clinical recovery. The study highlighted the need for further research, including randomized controlled trials to compare different doses and durations of corticosteroid therapy. It also emphasized the importance of monitoring patients for long-term complications, as well as the risks and benefits associated with hyperglycaemia and other side effects of corticosteroids.

In summary, the study found that corticosteroid therapy in COVID-19 pneumonia was associated with increased mortality and delayed clinical recovery when using pulse dose corticosteroids compared to conventional doses. However, it also demonstrated a significant reduction in inflammatory markers and improvements in various outcomes. The study’s findings contribute to the existing literature, with some unique aspects such as long-term follow-up and the need for further research to optimize corticosteroid therapy.

## Data Availability

All data produced in the present study are available upon reasonable request to the authors

